# Analysis of cell-specific peripheral blood biomarkers in severe allergic asthma identifies innate immune dysfunction

**DOI:** 10.1101/2022.01.12.22268892

**Authors:** Ben Nicholas, Jane Guo, Hyun-Hee Lee, Alistair Bailey, Rene de Waal Malefyt, Milenko Cicmil, Ratko Djukanovic

**Author notes:** **Correspondence to:** Dr Ben Nicholas, Centre for Proteomic Research, B85, Life Sciences Building, University of Southampton, University Road, Highfield, Southampton, Hants. SO17 1BJ.

## Abstract

Asthma is a disease of complex origin and multiple pathologies. There are currently very few biomarkers of proven utility in its diagnosis, management or response to treatment. Recent studies have identified multiple asthma phenotypes following biofluid analysis; however, such findings may be driven by the well-characterised alterations in immune cell populations in asthma. We present a study designed to identify cell type-specific gene signatures of severe allergic asthma in peripheral blood samples. Using transcriptomic profiling of four magnetically purified peripheral blood cell types, we identify significant gene expression changes in monocytes and NK cells but not T lymphocytes in severe asthmatics. Pathway analysis indicates dysfunction of immune cell regulation and bacterial suppression in the NK cells. These gene expression changes may be useful on their own as prognostic peripheral blood cell markers of severe asthma, but also may indicate novel cell pathways for therapeutic intervention.

---

Asthma is a common chronic airways disease with complex aetiology, having no single causative genetic trigger, and with multiple factors appearing to contribute to disease pathogenesis ^1^. Thus, asthma can manifest in numerous different pathologies related to severity and the degree of allergy or response to therapeutic intervention. Currently, the principal clinical biomarkers for initial diagnosis and staging of disease severity include lung function and hyper-responsiveness tests. Sometimes fractional exhaled nitric oxide (FeNO) is used as a surrogate for direct measurement of eosinophils. Improvement of clinical symptoms following therapeutic intervention and assessment of atopy and a history of wheeze are also useful.

Monitoring of the disease is generally through symptom assessment and therapeutic dose requirements combined with periodic lung function tests. Novel approaches, such as immunotherapy, have identified the utility of additional biomarkers such as IgE or sputum inflammatory cell counts as inclusion criteria; however, in clinical practice, often the principal read-outs remain the generalised clinical ones. No single test provides definitive evidence of asthma, its severity or therapeutic efficacy of drugs.

Recent reports identified multiple disease phenotypes using unbiased clustering of genomic and proteomic data from airway samples^2^. Likewise, mRNA signatures of severe asthma have been identified in whole blood ^3^. The importance of such studies may be confounded by known changes in inflammatory cell profiles in asthma, thus, missing changes in blood cell-subtypes by defining the phenotypic clusters of subjects based on inflammatory cell predominance in biofluids.

Our approach was to gene expression markers of cell type and asthma in four peripheral blood cell populations. We recruited 11 severe allergic asthmatics on step 4 of BTS/Sign management, and 10 healthy control subjects (see Table 1 for a summary of patient characteristics). Participants signed written informed consent.

**Table 1.** Clinical characteristics for the healthy and moderate/severe allergic asthmatic cohorts.

|  | Healthy controls | Severe allergic asthmatics | p value |
| --- | --- | --- | --- |
| N | 10 | 11 | - |
| Gender (M/F) | 3/7 | 3/8 | - |
| Age | 42.5 (31-53) | 43 (31-50) | 0.87 |
| Atopy (yes) | 0 | 11 | - |
| Total blood IgE | 16.45 (10.7-27.8) | 163.9 (71.4-315.0) | 0.0003*** |
| Blood lymphocytes | 1.9 (1.75-2.125) | 1.9 (1.7-2.1) | 0.345 |
| Blood eosinophils (%) | 0.1 (0.1-0.2) | 0.2 (0.1-0.3) | 0.212 |
| Pre FEV1 | 3.11 (2.89-4.13) | 2.58 (2.15-3.49) | 0.043* |
| Pre FEV1 % predicted | 111.4 (105.3-117.1) | 100 (77.2-102.4) | 0.016* |
| Post FEV1 | 3.19 (3.04-4.19) | 3.00 (2.37-3.66) | 0.067 |
| Post FEV1 % predicted | 114.1 (107.7-119.4) | 100.6 (86.2-114.7) | 0.055 |
| Sputum eosinophils (%) | 0 | 1.69 (0-3.68) | 0.046* |
| Sputum neutrophils (%) | 11.36 (7.1-15.7) | 64.46 (21.9-74.2) | 0.006** |
| PBMC CD4+ (% of CD3+) | 65.9 (53.5-71.2) | 70.3 (58.5-78.5) | 0.251 |
| PBMC CD8+ (% of CD3+) | 26.7 (18.6-33.0) | 23.9 (16.8-26.5) | 0.340 |
| PBMC monocytes (% of CD45+) | 11.7 (10.2-16.4) | 12.8 (8.7-14.0) | 0.878 |
| PBMC NK cells (% of Cd45+) | 11.1 (8.8-19.5) | 10.6 (7.4-11.7) | 0.314 |

Mann Whitney tests, \*p<0.05, \*\*p<0.01, \*\*\*p<0.001 Data shown are medians (+/- interquartile range)

Fresh blood was drawn and PBMCs were isolated from buffy coats. Flow cytometry determined no significant differences in the relative proportions of four immune cell populations, monocytes, NK cells, CD4+ and CD8+ T cells in the peripheral blood of healthy and severe asthmatic subjects (Table 1).

Four cell populations were sequentially enriched from 4 × 10^7^ PBMCs from each subject using magnetic beads conjugated to monoclonal antibodies targeting different cell surface markers, namely CD14 (monocytes), CD56 (NK/NK-T cells), CD8 (CD8+ T cells), and CD4 (CD4+ T cells) in that order. This yielded populations highly enriched (>99%) for each selective marker.

Quantification of gene expression used isolated RNA from the four enriched cell populations, analysed by a 770 gene nCounter pan cancer immune profiling array (NanoString Technologies).

Unbiased principal component analysis (PCA) of the purified cell gene expression profiles robustly separated the four target cell populations (Figure 1A). The genes primarily driving cluster separation of each cell type, and as such indicative of cell-specific biomarkers, confirm the purity of our immune blood cell populations and add to the canon of knowledge on markers of these cell types (see online data supplement for cell-specific gene lists: https://doi.org/10.6084/m9.figshare.19329068).

**Figure 1.**
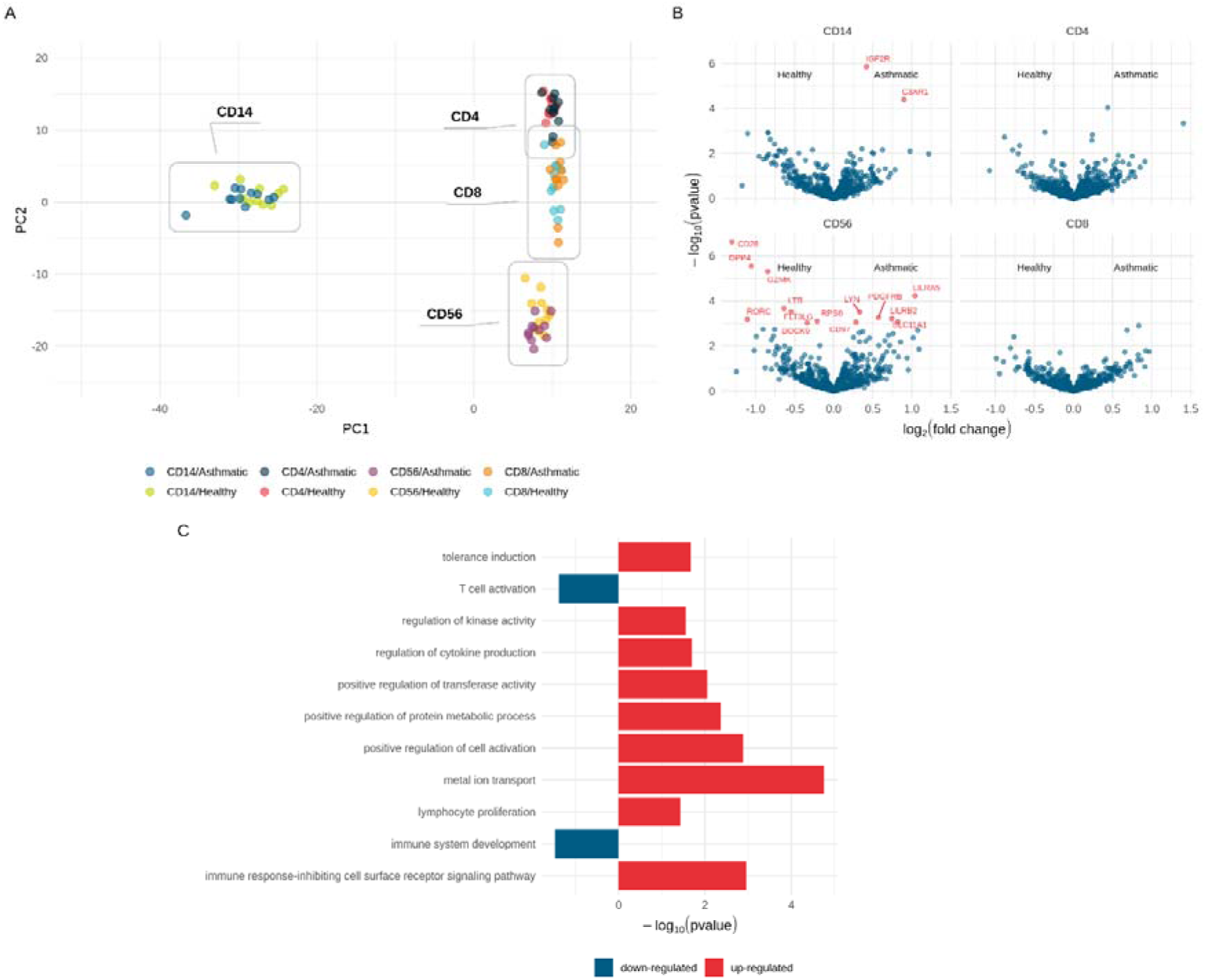
Differential gene expression analysis in RNA transcripts in severe asthma. **(A)** PCA analysis of gene counts from all four cell types indicate clusters corresponding to each cell type. **(B)** Volcano plots indicating genes significantly up or down regulated in severe asthma in each cell type (p-adjusted < 0.05). **(C)** Summary of GO terms associated with differential gene expression in NK cells in asthma.

For example, a major component of the monocyte cluster was CD14, which was to be expected since it was the target isolation molecule, but also includes 425 other genes contributing to this cluster, inclusive of the well-characterised monocyte-specific proteins such as CD68, CD163, L-selectin and S100A12.

T cells exhibit a well-defined cluster, however a small degree of overlap between the CD4+ and CD8 + T cell clusters, reflects the phenotypic similarity between these two cell types. Their combined cluster includes CD3 and CD8.

Similarly, our top-ranked genes contributing to the NK cell cluster are well-known phenotypic markers of NK cells, inclusive of KLRD1, NCAM-1 and EOMES. Clusters were driven primarily by cell type. We found little evidence of segregation between health and asthma in these cell populations.

We then identified differentially expressed genes (DEGs) in asthma, adjusting for multiple tests for each cell type as appropriate. Volcano plots (Figure 1B) indicate two genes upregulated in asthmatic monocytes compared to healthy controls (IGF2R and C3AR1). IGF2R is upregulated in monocytes during differentiation into macrophages ^4^ but changes in IGF2R expression by monocytes in asthma have not previously been observed. C3AR1 has been previously identified as a susceptibility gene for bronchial asthma ^5^. To our knowledge, no previous studies have identified elevation of the C3AR1 in monocytes of severe asthmatics.

We found six upregulated (LILRA5, LILRB2 and SLC11A1, PDGFRB, LYN, CD97) and eight downregulated (LTB, RORC, GZMK, DPP4, RPS6, FLT3LG, DOCK9 and CD28) genes in CD56+ cells from asthmatics.

Several of these upregulated genes are associated with immune cell regulation. For example, the leukocyte immunoglobulin-like immunoregulatory checkpoint receptors LILRA5 and LILRB2 have both positive and negative regulatory properties, whereas LYN-dependent signalling is primarily a negative regulator of innate and adaptive immune responses^6^. Additionally, the PDGF pathway has a well described local airways role in asthma pathogenesis and airway remodelling. Increased systemic levels of PDGF ligands in the serum of asthmatics have been previously observed, however it has remained uncertain whether PDGF is a systemic biomarker of asthma^7^. Our study suggests that PDGF-receptor expression may provide evidence of systemic response to abnormal PDGF expression in asthma.

Another upregulated gene in asthmatic NK cells is involved in pathogen resistance; SLC11A1 plays a role in divalent metal ion transport, promoting anti-microbial pathogen responses. This protein is upregulated in local innate lymphocytic cell subtypes in asthma, augmenting their activation and biasing towards a Th1 rather than Th2 response. Abnormal SLC11A1 expression has previously been identified in asthmatic sputum cells^8^, attributed to localised bacterial exposure. Systemic changes in expression of SLC11A1 may reflect the disease severity and the consequent neutrophilic phenotype of our severe asthma cohort. However, the existence of SLC11A1 upregulation in the peripheral is suggestive of widespread systemic innate cell dysfunction.

Conversely, several genes downregulated in CD56+ cells of asthmatics (LTB, RORC, GZMK, DPP4, RPS6, FLT3LG, DOCK9 and CD28), are associated with proinflammatory, functional, cell or activation responses in NK cells. For example, lymphotoxin beta is a pro-inflammatory TNF-α superfamily member, granzyme K is a serine protease and component of cytotoxic granules in NK cells, and CD28 is a co-stimulatory molecule for NK cells. Interestingly, DPP4, which plays a role in NK cell activation ^9^ is one of the ligands of IGF2R which is elevated in monocytes and may be suggestive of common systemic pathway alterations in peripheral innate cells.

Pathway analysis using the differentially expressed genes in the NK cell-enriched population (Figure 1C) confirmed that severe allergic asthma was associated with upregulation of pathways involved in metabolism, divalent metal ion transport, immune response inhibition, and immune regulation/tolerance, and with downregulation of T cell activation and immune system development.

Overall, these data suggest that an increased tolerogenic environment accompanies severe asthma, and NK cell dysfunctions that would impair the adaptive immune response were observed that could reflect exhaustion or generalised impairment of these cells. In addition, they show enhanced anti-microbial responses in the absence of recent exacerbation. Although NK and NK-like cells have been examined in asthma, no fixed role for them has yet been assigned. Our study suggests that further investigation of these cells is warranted.

Additionally, monocytes in asthma show increases in gene expression associated with differentiation and chemotaxis. Thus, we find innate but not adaptive cell phenotypic changes in peripheral blood cells from severe asthmatics.

Given the increasing interest in innate immunity and its role in antimicrobial responses, these results suggest the value of further study of gene expression changes at times of acute asthma exacerbations.

The findings presented herein may help to identify new therapeutic avenues for severe asthma arising from innate immunity, and also may serve to identify new gene candidates as useful cell type-specific biomarkers.

## Data Availability

All data produced in the present work are contained in the manuscript

